# Effect of long-term low dose glucocorticoid on children with bronchial asthma

**DOI:** 10.1101/2021.09.28.21264243

**Authors:** Xueping Du, Yanjun Guo, Junzheng Yang

## Abstract

**Objectives:** To investigate the clinical efficacy of long-term of low-dose inhaled glucocorticoid (ICS) in the treatment on children with bronchial asthma, and to evaluate its effects on the height, weight and expression of serum insulin-like growth factor of children with bronchial asthma.

**Methods:** 91 children with bronchial asthma treated in our hospital from January 2017 to December 2017 were chosen, and 31 healthy children without asthma history in our hospital were selected as the control group (Group C); For the different treatment methods comparison, 91 children with bronchial asthma were divided into treatment group A (48 children) and treatment group B (43 children) randomly. In treatment group A, children were treated with low-dose ICS for more than 1 year; in treatment group B, children were treated with low-dose ICS for less than 1 year or received low-dose ICS for less than 2 months/year. For treatment evaluation, the asthma control test (C-ACT) were scored at 0 months, 12 months and 24 months in three groups, and the height and weight of children, the levels of insulin-like growth factor-1 (IGF-1) and insulin-like growth factor binding protein-3 (IGFBP-3) in serum in each group were also measured.

**Results:** In the control group, the C-ACT scores were increased with time (P<0.01); the C-ACT scores of treatment group A at 12 months and at 24 months was significantly higher than that of treatment group B (P<0.05); there was no significant difference in height and weight between the treatment group and the control group at months, 12 months and 24 months after treatment (P>0.05); there was no significant difference in the expression levels of IGF-1 and IGFBP-3 in serum between treatment group and control group (P>0.05).

Compared with the C-ACT scores in treatment group A and treatment group B at 0 month, the C-ACT scores were significantly higher at 12 months and at 24 months, and there was a statistical difference among 0 month, 12 months and at 24 months in treatment group A and treatment group B (P<0.01); The C-ACT score of treatment group A at the 12 month was significantly higher than that in treatment group B at the 12 months(P<0.05), and the C-ACT score in treatment group A at the 24 months was significantly higher than that of treatment group B (P<0.001).

**Conclusions:** Treatment for children with bronchial asthma by long-term use of low-dose ICS is safe and effective, it does not affect health and development of children with bronchial asthma; children with bronchial asthma were treated with low-dose ICS for more than 1 year had a better effective than children were treated with low-dose ICS for less than 1 year or received low-dose ICS for less than 2 months/year.

## Introduction

Bronchial asthma is a common respiratory disease characterized by chronic airway inflammation and airway hyperresponsiveness^[1]^. Long-term use of inhaled corticosteroids (ICS) is the main means to treat asthma. It can control airway inflammation, reduce airway hyperresponsiveness, asthma symptoms, the number of acute attacks of asthma and the mortality of severe asthma^[2]^. Although long-term use of ICS has significant clinical efficacy, parents worry that long-term use of ICS will affect children’s development, especially height and weight, indirectly leading to unsatisfactory treatment effect of children with asthma and reduction of quality of life^[3]^. Therefore, it is of great clinical and social significance to study the effect of long-term use of low-dose ICS on children with asthma and its impact on height and weight. It is found that the levels of serum insulin-like growth factors-1 (IGF-1) and insulin-like growth factor binding protein-3 (IGFBP-3) are mainly regulated by growth hormone, which can reflect the state of growth hormone. The detection of IGF-1 and IGFBP-3 in serum reflects the state of growth and development at the biochemical level^[4]^, the effect of long-term inhalation of low-dose ICS on IGF secretion in children bronchial asthma is not very clear. For above purpose, 91 children with bronchial asthma treated in our hospital were given long-term low-dose ICS, and the asthma control test score^[5]^, height and weight, expression of IGF-1 and IGFBP-3 in children were measured, to investigate the effectiveness of long-term use of ICS in the treatment of bronchial asthma and its impact on children’s growth and development.

## 1 Investigation subjects and methods

### 1.1 Investigation subjects

91 children with bronchial asthma treated in our hospital from January 2017 to December 2017 were collected according to GINA asthma diagnostic criteria in 2016 ^[6]^, all the children (aged 4-9 years old) were treated by standard treatment of long-term use of low-dose ICS. In treatment group A, children were treated with low-dose ICS for more than 1 year, in treatment group B, children were treated with low-dose ICS for less than 1 year or received low-dose ICS for less than 2 months/year; and 31 healthy children without asthma history in our hospital were selected as the control group (Group C). All enrolled children were excluded from the following situations: (1) chronic diseases (liver, kidney, endocrine and metabolic system, all kinds of congenital heart diseases, immune deficiency diseases, bone development diseases); (2) children were in puberty, low birth weight children, whose weight is 2 standard deviations lower than that of normal children in the same age; (3) receiving other types of glucocorticoids previously. There were 47 boys and 44 girls in the treatment group and 16 boys and 15 girls in the control group; There was no difference in age, sex, height and weight between the two groups (P>0.05).

### 1.2 Investigation methods

Children in the treatment group inhaled low-dose fluticasone propionate (trade name: fluticasone, specification: 125μg×60 press, produced by GlaxoSmithKline Pharmaceutical Co., Ltd.) or salmeterol/fluticasone (trade name: Seretide, specification: 50μg/100μg×60 bubbles/each box, produced by GlaxoSmithKline Pharmaceutical Co., Ltd.) as basic maintenance treatment, asthma outpatient nurses taught children and parents how to use it. Low dose was defined as the daily average dose of fluticasone≤200μ g/d. Other adjuvant drugs were determined according to age, attack degree and attack frequency according to GINA guidelines in 2016; The same pediatrician would visit the asthma clinic of our hospital every 3 months to evaluate the asthma control of children.

### 1.3 Detection of evaluation indexes

C-ACT test were performed in children with bronchial asthma at 0 month, 12 months and 24 months after treatment; the height and weight of children in the three groups were measured and recorded by professionally trained doctors. For expression of IGF-1 and IGFBP-3, 2 mL of femoral vein blood on an empty stomach at 0 month, 12 months and 24 months after treatment was taken, separated the serum, and detected the expression of IGF-1 and IGFBP-3 in the serum of children in each group by ELISA, the operation was carried out in strict accordance with the instructions of the kit.

### 1.4 C-ACT score

C-ACT score was studied and published by the National Jewish Medical Research Center of the United States. It can reflect the control state of asthma in clinical application, and is also used to monitor the condition of children’s asthma and evaluate the clinical efficacy of asthma drug treatment^[7]^. C-ACT Score is a questionnaire to evaluate asthma control according to the clinical symptoms of children in recent 4 weeks. The full score is 27 points, the score is lower than 19 points defined as uncontrolled, 20-24 points are defined as basic control, and 25-27 points are defined as complete control.

### 1.5 Statistical analysis

The data were analyzed by SPSS 23.0 software. ANOVA analysis were used to compared the difference among three groups, the data were showed as mean±standard deviation 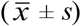. *P*<0.05 indicated that there was a statistically difference.

## 2. Results

### 2.1 Comparison of height and weight of children with bronchial asthma between the treatment group and the control group at 0 month, 12 months and 24 months before and after treatment

Compared with 0 month after treatment, the height and weight of children in the treatment group or in the control group were increased significantly at 12 and 24 months (P<0.01); there was no significant difference between treatment group and control group at 0 month, 12 months and 24 months after treatment (P>0.05). Those data demonstrated that there was no effect of the long-term use of low-dose ICS on weight or height of children with bronchial asthma (Table 1).

**Table 1.** Comparison of height and weight of children with bronchial asthma between the Treatment group and the control group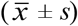

| Treatment group and the control group($\bar{x} \pm s$) | | | | | | | |
| --- | --- | --- | --- | --- | --- | --- | --- |
| Group | n | Height (cm) |  |  | Weight(kg) |  |  |
|  |  | 0 month | 12 months | 24 months | 0 month | 12 months | 24 months |
| Treatment group | 91 | 113.95±10.64 | 120.82±10.95 | 128.78±11.75 | 18.95±3.29 | 21.57±3.69 | 24.82±4.09 |
| Control group | 31 | 116.14±12.11 | 123.78±12.59 | 131.93±13.52 | 19.63±3.26 | 22.27±3.53 | 25.74±4.07 |
| 1 | | F=2458.74 | | $p<0.01$ | | F=1718.58 | $p<0.01$ |
| 2 | | F=2.69 | | $p>0.05$ | | F=0.65 | $p>0.05$ |
| 3 | | F=1.34 | | $p>0.05$ | | F=1.02 | $p>0.05$ |

1: Comparison between 0 month, 12 months and 24 months after treatment in treatment group or in control group;
2: Comparison between treatment group and control group at 0 month, 12 months and 24 months after treatment;
3: Comparison between treatment group and control group.

### 2.2 Comparison of the expression of IGF-1 and IGFBP-3 between the treatment group and control group before and after treatment

Compared with 0 month after treatment, there were a statistical difference in expression of IGF-1 in serum of children at 12 months and 24 months in treatment group (P<0.05) but not control group (P>0.05); compared with 0 month after treatment, there were no statistical difference in expression IGFBF-3 in serum of children at 12 months or 24 months in treatment group and control group (P>0.05); compared with the control group, there was no statistical difference in the expression of IGF-1 or IGFBF-3 in serum at 0 month, 12 months and 24 months in the treatment group (P>0.05) (Table 2). Those data demonstrated that long-term use of long-term use of low-dose ICS had no effect on the expression of IGF-1 and IGFBP-3 in children with bronchial asthma (Table 2).

**Table 2.** Comparison the expression of IGF-1 and IGFBP-3 in children with bronchial asthma between treatment group and control group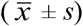

| group | n | IGF-1(ng/mL) |  |  | IGFBP-3 (ng/mL) |  |  |
| --- | --- | --- | --- | --- | --- | --- | --- |
|  |  | 0 month | 12 months | 24 months | 0 month | 12 months | 24 months |
| Treatment group | 91 | 121.47±15.63 | 127.54±13.44 | 128.89±13.86 | 43.28±9.41 | 43.97±10.27 | 43.00±9.76 |
| Control group | 31 | 125.92±16.86 | 127.62±10.56 | 129.92±9.78 | 44.50±8.95 | 45.71±8.56 | 47.49±9.48 |
| 1 |  |  | F=8.30 p<0.05 |  |  | F=1.40 p>0.05 |  |
| 2 |  |  | F=1.30 p>0.05 |  |  | F=2.25 p>0.05 |  |
| 3 |  |  | F=0.60 p>0.05 |  |  | F=2.00 p>0.05 |  |

1: Comparison between 0 month, 12 months and 24 months after treatment in treatment group or in control group ;
2: Comparison between treatment group and control group at 0 month, 12 months and 24 months after treatment;
3: Comparison between treatment group and control group.

### 2.3 Comparison of C-ACT scores in 91 cases children with bronchial asthma at 0 month, 12 months and 24 months after treatment

The results of C-ACT showed that C-ACT scores of children with bronchial asthma in the treatment group at 12 months and 24 months were higher compared that in 0 month, and there was a statistical difference at 0 month, 12 months and 24 months in treatment group (P<0.05) (Table 3). The results showed that long-term use of low-dose ICS could alleviated the symptoms of children with bronchial asthma effectively (Table 3).

**Table 3.** Comparison of the C-ACT scores before at 0 month, 12 months and 24 months in 91 cases children with bronchial asthma

| Group | Cases | Score |  |  |
| --- | --- | --- | --- | --- |
|  |  | 0 month | 12 months | 24 months |
| Treatment group | 91 | 16.28±3.06 | 20.65±2.43 | 22.88±2.86* |
| F=149.69, p<0.05 |  |  |  |  |

### 2.4 Comparison of C-ACT scores in treatment group A and treatment group B

Compared with the C-ACT scores in treatment group A and treatment group B at 0 month, the C-ACT scores were significantly higher at 12 months and at 24 months, and there was a statistical difference among 0 month, 12 months and at 24 months in treatment group A and treatment group B (P<0.01); The C-ACT score of treatment group A at the 12 month was significantly higher than that in treatment group B at the 12 months(P<0.05), and the C-ACT score in treatment group A at the 24 months was significantly higher than that of treatment group B (P<0.001). the results indicated that children with bronchial asthma were treated with low-dose ICS for more than 1 year (treatment group A) had a better effective than children were treated with low-dose ICS for less than 1 year or received low-dose ICS for less than 2 months/year (treatment group B)(Table 4).

**Table 4.** Comparison of C-ACT scores of treatment group A and treatment group B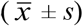

| Group | cases | Score |  |  |
| --- | --- | --- | --- | --- |
|  |  | 0 month | 12 months | 24 months |
| Treatment group A | 48 | 16.33±3.23 | 21.13±2.39 | 24.00±2.32 |
| Treatment group B | 43 | 16.16±2.86 | 20.07±2.38 | 21.63±2.93 |
| 1 | | | F=154.49 $p<0.01$ | |
| 2 | | | F=4.25 $p<0.05$ | |
| 3 | | | F=11.05 $p<0.05$ | |

1: Comparison between 0 month, 12 months and 24 months after treatment in treatment group A or in treatment group B; 2: Comparison between treatment group A and in treatment group B at 0 month, 12 months and 24 months after treatment; 3: Comparison between treatment group A or in treatment group B.

## 3. Discussion

Most children with bronchial asthma began to appear symptoms before 3 years old, the early symptoms are not obvious, which is not easy to attract the attention of parents. If timely treatment is not taken, it will easily lead to delayed healing and impaired pulmonary function, which will increase the difficulty of treatment and seriously affect the quality of life of children^[8]^. Bronchial asthma is characterized by chronic airway inflammation, the pathophysiological process is mainly airway hyperresponsiveness and airway remodeling^[9]^, and glucocorticoid is the most effective anti-allergic inflammatory drug at present, which can significantly inhibit airway inflammation. ICS is a local drug in the airway, which can not only control the acute attack of bronchial asthma^[10]^, improve peak expiratory flow rate, inhibit airway hyperresponsiveness and prevent seizures, can also reduce its side effects. It is the first choice for the long-term treatment of children with bronchial asthma^[11]^. Children with bronchial asthma usually need long, continuous and standardized inhalation treatment to achieve the expected effect. It is recommended that the continuous inhalation time be initially set at 6 months to 1 year. If bronchial asthma symptoms relapse, ICS should be used again according to the intensity and frequency of bronchial asthma attack^[12]^.

In this study, we found that standardized inhalation of low-dose fluticasone propionate or salmeterol/fluticasone for 12 and 24 months, the C-ACT score of children with bronchial asthma was significantly higher than that at 0 month, and the score of patients at 24 months was higher than that at 12 months, those data demonstrated that long-term continuous standardized inhalation of low-dose fluticasone propionate or salmeterol/fluticasone was effective for bronchial asthma treatment with the treatment time. In addition, the C-ACT score of treatment group A at the 12th month was significantly higher than that in treatment group B, and the C-ACT score increased after 24 months. It is speculated that children in treatment group A have standardized inhalation of low-dose ICS for less that one year, and the compliance is good, while children in treatment group B have treated with low-dose ICS for less than 1 year or have previously received inhalation ICS for no more than 2 months/year, and the compliance is general, indicating that children with good compliance have better bronchial asthma control. Therefore, long-term continuous use of ICS with good compliance are more conducive to the control of bronchial asthma in children. It is suggested that in the process of managing children with bronchial asthma, the medical staff should strengthen the management of standardized use of low-dose ICS treatment for children with bronchial asthma, and take individualized intervention measures to improve the compliance of children with bronchial asthma with ICS treatment, so as to achieve the purpose of standardized management is to improve the pulmonary function and airway hyperresponsiveness of asthmatic children, improve the quality of life of asthmatic children, and achieve the purpose of completely controlling bronchial asthma^[13]^.

Although ICS is the most effective drug for the treatment of bronchial asthma, its effect on children’s growth and development is still controversial^[14]^. Long-term intravenous or oral glucocorticoid treatment of bronchial asthma is easy to cause fever, osteoporosis, obesity and decreased osteoblast activity in children, resulting in abnormal growth and development^[15,16]^. In order to study whether the long-term use of ICS has an impact on the growth and development of children, we recorded the height and weight of children in each group. the results demonstrated that the height and weight of children in the treatment group and the control group were in line with the general law of height and weight growth of normal children, and there was no significant difference between the treatment group and control group, It demonstrated that the long-term use of ICS for treatment of asthma does not affect the height and weight of children. Children’s growth is affected by congenital heredity, acquired nutrition, environment and medication history, and is jointly regulated by growth hormone (GH)-IGFs IGFBPs system. The expression of IGF-1 and IGFBP-3 are the key markers for reflecting the changes of growth hormone insulin-like growth factor axis^[17,18]^. IGF-1 can promote the growth of whole body organs, promote the differentiation and proliferation of chondrocytes in epiphyseal plate, and play an important role in bone development. IGFBPs are specific proteins with high affinity with IGFs, which have the highest content in blood and are specific for detecting abnormal height in children^[19]^. Our study found that there was no significant difference IGF-1 and IGFBP-3 levels in serum between treatment group and control group after 12 and 24 months of treatment with ICS, indicating that long-term use of ICS had no effect on GH-IGFs-IGFBPs system, it is proved that the long-term low-dose use of ICS will not affect the growth and development of children. In order to better evaluate the impact of long-term use of ICS on children’s growth and development, longer follow-up is necessary.

In summary, the long-term standardized use of low-dose ICS for children bronchial asthma treatment had obvious clinical efficacy and high safety, and d id not affect the growth and development of children.

## Data Availability

The data used to support the findings of this study are available from the corresponding author upon request.

## 4. Conflicts of Interest

The authors declare they have no conflicting financial interests.

## 5. Funding Information

None.

## 6. acknowledgements

None.

## Notes

### Competing Interest Statement

The authors have declared no competing interest.

### Author Declarations

I have the right to post this manuscript and confirm that all authors have assented to posting of the manuscript. All relevant ethical guidelines have been followed. Details of the oversight body: Ethics committee of Children's Hospital Affiliated to Zhengzhou University suggested that Effect of long-term low dose glucocorticoid on children with bronchial asthma is ethical in according with Ethics committee of Children's Hospital Affiliated to Zhengzhou University, and agree to conduct relevant experiments. All necessary patient/participant consent has been obtained. I am legally responsible for the content of the article. I have followed all appropriate research reporting guidelines

## References

[1] Papia, Brightling C, Pedersen SE, et al. Asthma [J]. Lancet, 2018, 391(391): 783–800.

[2] Boulet LP, Fitzgerald JM, Reddel HK. The revised 2014 GINA strategy report: opportunities for change [J]. Curr Opin Pulm Med, 2015, 21(21): 1–7.

[3] Respiratory group, scientific branch, Chinese Medical Association. Guidelines for diagnosis, prevention and treatment of bronchial asthma in children (2016 Edition) [J]. Chinese Journal of Pediatrics, 2016, 3 (3): 167-181 (In Chinese).

[4] Weifu Chen, Aiping Zhou, Hongdan Gu, et al. The value of serum insulin-like growth factor-1, insulin-like growth factor binding protein-3 and bone age in the diagnosis of children with dwarfism [J]. China maternal and child health care, 2018, 09): 2047-2050 (In Chinese).

[5] Zhiping Li, Lihong Peng, yubiao Guo, et al. Feasibility study on the application of asthma control test in China [J]. China Medical Engineering, 2007, 2 (2): 160-162 (In Chinese).

[6] Global Initiative for Asthma. GINA Report: Global Strategy for Asthma Management and Prevention (2016). Available online at: https://ginasthma.org/gina-reports/.

[7] Yimin Yang, Gulan Zeng, Yaxin Li, et al. Clinical application of bronchial asthma control test in children [J]. Chinese Journal of Practical Pediatrics, 2017, 16 (16): 1248-1252 (In Chinese).

[8] Ducharme FM, Tse SM, Chauhan B. Diagnosis, management, and prognosis of preschool wheeze [J]. Lancet, 2014, 383(383): 1593–604.

[9] Xiaoguang Hu, Hongmei Yu, Qiuping Wu, et al. Prognostic follow-up of recurrent wheezing in infants and analysis of risk factors for persistent wheezing [J]. Journal of Wenzhou Medical University, 2016, 5 (5): 365-368(In Chinese).

[10] Yingchun Yang. Clinical study on the effectiveness of aerosol inhalation of budesonide and oral prednisone on the acute attack of cough and asthma in children [J]. Journal of medical forum, 2019, 40 (40): 31-34 (In Chinese).

[11] Bush A. Management of asthma in children [J]. Minerva Pediatr., 2018, 70(70): 444–457.

[12] Yanhan Zhang, Yuan Zhou, Yanming Lu. Research progress on treatment compliance of inhaled corticosteroids in children with asthma [J]. Chinese Journal of lung diseases (Electronic Edition), 2019, 3 (3): 372-374 (In Chinese)..

[13] Geryk LL, Roberts CA, Carpenter DM. A systematic review of school-based interventions that include inhaler technique education [J]. Respir Med, 2017, 132:21–30.

[14] Loke YK, Blanco P, Thavarajah M, et al. Impact of Inhaled Corticosteroids on Growth in Children with Asthma: Systematic Review and Meta-Analysis [J]. PLoS One, 2015, 10(10): e0133428.

[15] Ramadan A A, Gaffin JM, Israel E, et al. Asthma and Corticosteroid Responses in Childhood and Adult Asthma [J]. Clin Chest Med, 2019, 40(40): 163–177.

[16] Shengye Liu, Qin Fu. Latest research progress on the pathogenesis of glucocorticoid induced osteoporosis [J]. Chinese Journal of bone and joint, 2015, 11:877-880 (In Chinese).

[17] Renes JS, Van Doorn J, Hokken-Koelega ACS. Current Insights into the Role of the Growth Hormone-Insulin-Like Growth Factor System in Short Children Born Small for Gestational Age [J]. Horm Res Paediatr, 2019, 92(92): 15–27.

[18] Blum WF, Alerbish A, Alsagheir A, et al. The growth hormone-insulin-like growth factor-I axis in the diagnosis and treatment of growth disorders [J]. Endocr Connect, 2018, 7(7): R212–R22.

[19] Ranke MB. Insulin-like growth factor binding-protein-3 (IGFBP-3) [J]. Best Pract Res Clin Endocrinol Metab, 2015, 29(29): 701–711.

